# Health, Equity, and Economic Impacts of a Nicotine Product Standard in the United States for People With and Without Major Depression

**DOI:** 10.1101/2025.07.10.25331302

**Authors:** Sarah Skolnick, Andrew Brouwer, Chia-ling Cheng, Jamie Tam

## Abstract

**Importance:** The U.S. FDA has proposed a product standard that would reduce the nicotine content and addictiveness of cigarettes. It is unclear what impact this would have on economic outcomes or priority populations who are disproportionately harmed by tobacco use, such as people with major depression.

**Objective:** To evaluate the long-term health and economic impacts of a nicotine product standard for the U.S. population by depression status.

**Design, Setting, and Participants:** A microsimulation model was developed and calibrated to National Survey on Drug Use and Health (NSDUH) 2005-2023 data on smoking, vaping, and depressive episodes. The anticipated effects of the nicotine product standard on smoking and vaping were obtained from an FDA expert elicitation and used to simulate the policy from 2027-2100.

**Exposure:** Smoking and vaping.

**Main Outcome(s) and Measure(s):** Health outcomes included prevalence of smoking and vaping, deaths, and life years gained overall and by major depression status. Economic outcomes include direct costs to the healthcare system and societal costs.

**Results:** Under the proposed nicotine product standard, smoking is projected to decline to <1% for people with and without depression by 2100. The policy is estimated to avert 1.7 million premature deaths and lead to 74.7 million life years gained. Depression prevalence is also expected to decline, with 8.5 million fewer cases of depression estimated. Longer life expectancies under the policy are projected to increase medical costs by $296 billion, while also increasing worker productivity by $266 billion with an additional $1.2 trillion in consumer spending.

**Conclusions and Relevance:** Timely implementation of a nicotine reduction strategy, either through a federal product standard or state-level sales restrictions, is cost-effective and could prevent millions of premature deaths and reduce smoking disparities by depression status.

## Introduction

The FDA has proposed a product standard that would limit nicotine content in combustible tobacco products, including cigarettes, which are the leading cause of preventable death in the United States.^1–3^ ^4,5^ This regulation would greatly decrease the addictive potential of cigarettes because nicotine is the primary addictive component in cigarettes, increases progression from experimentation to regular use, and makes quitting smoking extremely difficult.^4,5^ Studies show that, relative to normal nicotine content cigarettes (NNC), very low nicotine cigarettes (VLNC) increase smoking abstinence and quit attempts,^4^ with fewer cigarettes smoked per day,^6^ and there is no evidence of compensatory smoking (i.e., smoking more VLNCs to obtain the desired amount of nicotine).^4^ The FDA anticipates that the nicotine product standard could bring dramatic reductions in smoking-related mortality, resulting in medical cost savings and productivity gains. However, the potential economic and health equity impacts of a nicotine product standard are unknown. Tobacco manufacturers argue that the FDA is likely substantially underestimating the potential costs of the policy to society and that a nicotine product standard could harm people with mental health conditions.^7,8^

Evidence from randomized controlled trials indicates that people with mental health conditions could benefit from a nicotine product standard: persons with affective disorders randomized to receive VLNCs experienced reduced smoking, cigarette cravings, and depressive symptoms.^4,9^ The latter finding is especially noteworthy because of the bidirectional relationship between smoking and depression^10–14^: Mendelian randomization studies have identified causal links between smoking and depression,^13,15^ and smoking cessation decreases depression symptoms and increases positive affect.^16^ Given rising levels of major depression (MD) among youth and young adults,^17^ it is critical to understand the potential mental health implications of proposed tobacco regulations. No study has quantified the potential impacts of a nicotine product standard for people with mental health conditions. The present study evaluates the proposed rule’s projected impact on the health of people with and without MD, as well as the expected changes to healthcare and societal costs.

## Methods

### Model Overview

We developed the MD, smoking, and e-cigarettes (MDSE) microsimulation model to analyze the health and economic outcomes of a nicotine product standard for the US populations with and without MD. We simulated 20,000 men and women for each birth cohort from 1900 to 2100, totaling 4 million in the modeled population. Individuals enter the model at birth having never smoked, vaped, or had a MD episode. They can then transition across health states defined by depression status (never MD, current MD, and recovered from MD), smoking (never smoking, current smoking, and former smoking), and vaping (never vaping, current vaping, former vaping). Figure 1 illustrates all 27 health states and transitions between them. We simulated the nicotine product standard starting in 2027, consistent with the FDA proposed rule, accumulating health and economic impacts from 2027–2100. We assessed smoking and vaping prevalence, premature deaths averted, and life years gained in people with and without MD. We also calculated health and non-healthcare costs and productivity for the overall population.

**Figure 1.**
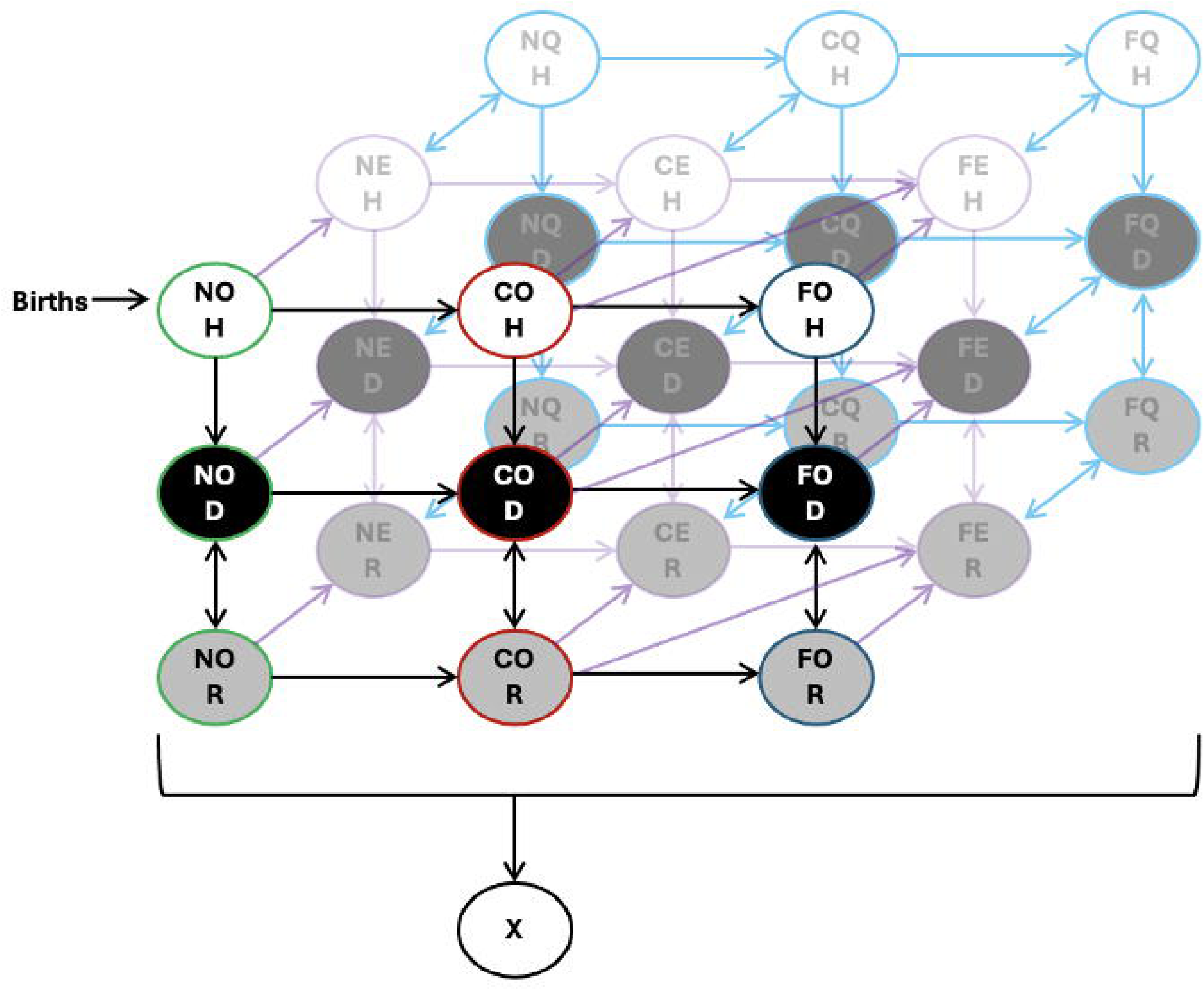
Overview of product use and depression status transitions in the Major Depression, Smoking, and E-cigarette Model (MDSE) Notes: N = Never smoking, C = Current smoking. F = Former smoking, O = Never vaping, E = Current vaping, Q = Former vaping = Never had major depression (Never MD), D = Depressed (Current MD), R = Recovered (Former MD), X = Dead.

### Data Sources

Transition probabilities between depression states were based on prior analysis of 2005 to 2023 National Surveys on Drug Use and Health (NSDUH) and the Baltimore Epidemiological Catchment Area data.^18^ Transitions between smoking states were based on the CISNET Lung Working Group estimates of annual smoking initiation and cessation.^19^ Transitions between smoking and vaping were based on analyses of the Population Assessment on Tobacco and Health.^20–22^ These probabilities were then calibrated to MD, smoking, and vaping prevalence by age and calendar year in NSDUH data (2005–23). Model parameters not available from the literature were directly estimated through calibration to NSDUH. For further details, see eMethods and eTable 1.

We used mortality probabilities, which vary by smoking status and years since quit, from the CISNET Lung Working Group .^19^ The exact mortality risk of e-cigarettes is unknown but is expected to be substantially lower than that of cigarettes. Although some public health entities estimate that vaping confers 5% of the mortality risk of smoking,^23,24^ we made a conservative assumption of 10% and varied the risk from 0–15% in sensitivity analysis.

### Nicotine Product Standard

We simulated a policy scenario using the FDA expert elicitation estimates of the effects of a nicotine product standard on smoking initiation, smoking cessation, dual use, product switching, and vaping initiation among those deterred from smoking.^25^ These effects were applied from 2027–2100. We also evaluated optimistic and pessimistic policy effect estimates based on the potential for the effect to facilitate migration away from or towards smoking; these reflect the 5^th^ and 95^th^ percentile values specified in the expert elicitation. See eMethods, eFigure 1, and eTable 4 for further details. In the absence of estimates by mental health status, we assumed these effects are the same for individuals with and without MD. Modeled outcomes are compared to a status quo scenario in which no nicotine product standard is implemented and existing tobacco use transitions continue unchanged.

### Product Use and Health Outcomes

We compared projected smoking and vaping prevalence for the U.S. population with MD and without MD. The absolute smoking disparity was calculated as the absolute difference in smoking prevalence between the two groups.

Premature deaths in the microsimulation model were scaled to the U.S. population by scaling modeled deaths in 2023 to actual U.S. deaths in 2023. Similarly, population life years were calculated by scaling up each birth cohort based on the number of actual births in the U.S. Premature deaths averted were calculated by subtracting the cumulative premature deaths of the policy scenario from the status quo scenario. Life Years Gained (LYG) were calculated as the difference in cumulative population life years between the status quo scenario and the policy scenario.

### Economic Evaluation

We estimated costs from both the healthcare and societal perspectives under the product standard and status quo scenarios. Healthcare costs per year were assigned to each individual by age, smoking status, and depression status based on data from the Medical Expenditure Panel Survey (MEPS).^26,27^ Societal costs include medical costs, consumer expenditures, and productivities. Consumption of non-healthcare goods (consumer expenditure) was assigned to each individual by age using the U.S. Bureau of Labor Statistics data.^28^ Individual productivities by age were based on total mean wages with fringe benefits (2023 fringe rate of 31%)^29^ and were estimated using the Current Population Survey, 2023 Annual Social and Economic Supplement (CPS ASEC).^30^ See eTable 3 for further cost details.

Utilities by age, smoking status, and depression status were calculated by converting the number of healthy days reported in the 2022–2023 Behavioral Risk Factor Surveillance System (BRFSS) data into EQ-5D scores (see eMethods).^31^ These utilities were used to calculate Quality Adjusted Life Years (QALYs) gained.

We calculated Incremental Cost Effectiveness Ratios (ICERs) as the cost difference between the policy and status quo scenarios divided by QALY gains under the policy. We then assessed the cost-effectiveness by comparison to a standard willingness-to-pay threshold of $100,000.^32,33^ Although, not standard, we also calculated ICERS for LYG and used it to obtain our estimates for U.S.-level costs. Our simulated population was smaller than the true U.S. population, and thus, to calculate costs at the population level, ICERs for each cost category were multiplied by discounted LYG (scaled to the U.S. population). We adhered to recommendations from the Second Panel on Cost-Effectiveness in Health and Medicine by including productivity costs in the numerator of the ICER,^34^ but we acknowledge that this approach is an area of ongoing debate.^35^ Note that we quantified QALYs using the EQ-5D, which is relatively insensitive to changes in income.^34,36^ Costs were inflated to 2023 dollars and discounted at 3% for projections after 2025. LYG and deaths averted were similarly discounted.

### Sensitivity Analyses

We conducted three sets of deterministic one-way sensitivity analyses to evaluate the influence of model parameters with the most uncertainty:

1. *Effects of nicotine product standard on product use transitions.* We separately applied each nicotine product standard policy effect at a time and calculated the cumulative premature deaths averted for each scenario. We also compared these mortality reductions to a best-case scenario: the Maximum Potential Reduction in Premature Mortality (MPRPM) scenario^37^ assumes all individuals quit smoking and no one initiates smoking from 2027–2100. Policy effect parameters that achieve larger shares of the MPRPM have greater influence on model outcomes compared to those that achieve smaller shares of the MPRPM.
2. *Excess mortality risk associated with vaping.* We compared e-cigarette mortality effects from 0–15% of the excess mortality risk associated with current smoking.
3. *Risk of developing MD.* To understand how our results may be influenced by increasing risk of developing MD since 2016, we also modeled a scenario where the risk of developing MD returned to its pre-2016 values starting from 2023 onwards.

## Results

### Product Use and Health Outcomes

The model aligned well with smoking, vaping, and dual use prevalence and trends overall and by MD status in the NSDUH data (Figure 2). Smoking prevalence decreased for all populations over time under the status quo, but more dramatic decreases occurred under a nicotine product standard, with smoking prevalence reaching <1% for all groups by 2040 (Table 1). Under the product standard, overall smoking prevalence reached 0.1% (0.0%, 0.6%) in 2100 compared to 3.2% under the status quo. By 2100, the absolute difference in prevalence between those with and without current MD decreased from 4.7% under the status quo to 0.5% under the nicotine product standard scenario. Prevalence dropped to 0.5% for those with MD and ∼0.0% for those without MD; at zero and near-zero prevalence, relative differences may appear disproportionately high and artificially suggest greater disparities. Following a nicotine standard, the model projected an initial increase in vaping initiation, as those deterred from smoking take up vaping instead. However, vaping was then projected to decrease as smoking declines, eventually plateauing over subsequent decades. MD prevalence was projected to decrease slightly under a nicotine product standard, reflecting the effects of reduced smoking on depression incidence and recurrence (see eFigure 2).

**Figure 2.**
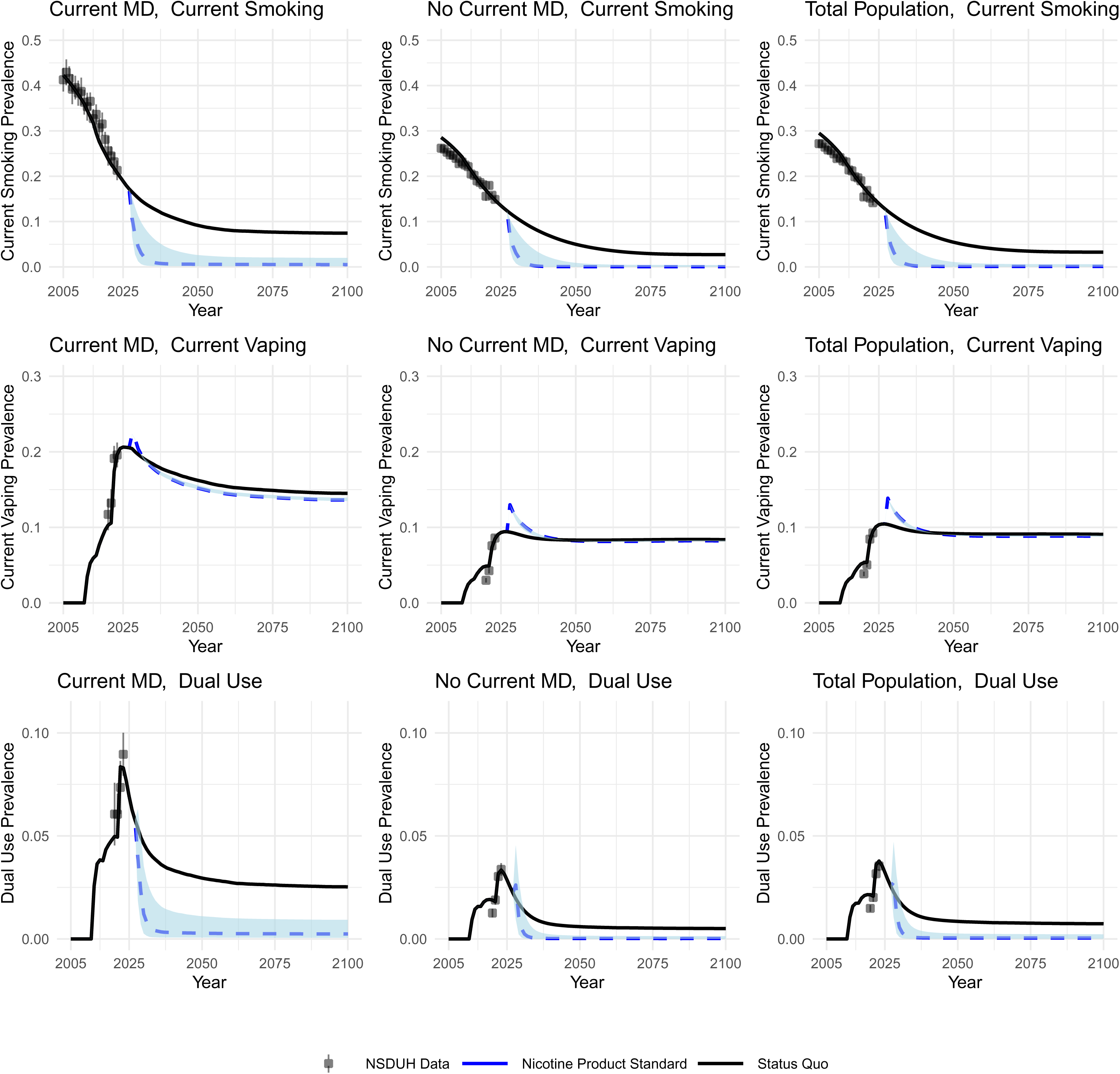
Actual and projected Smoking and Vaping Prevalence 2005-2100 by Major Depression (MD) Status in status quo and nicotine product standard (policy) scenarios Notes: The black solid line represents the status quo scenario, and the blue dashed line is the Nicotine Product Standard scenario. Light blue shading represents the area covered by the upper and lower bounds of optimistic and pessimistic scenarios. Dots are NSDUH data points used to calibrate the model. Vertical lines on dots are confidence intervals. Note that intervals are not visible for total and Never MD populations because large sample sizes resulted in small confidence bounds.

**Table 1.** Current and simulated Smoking Prevalence, 2026-2100, by Major Depression Status in status quo and nicotine product standard (policy) scenarios.

| Current Smoking Prevalence |  |  |  |  |  |  | Absolute Difference between those with and without MD |  |
| --- | --- | --- | --- | --- | --- | --- | --- | --- |
| Overall Population |  |  | Population with MD |  | Population without MD |  |  |  |
| Year | Status Quo | Policy | Status Quo | Policy | Status Quo | Policy | Status Quo | Policy |
| 2026 | 12.8% | 12.8% | 17.2% | 17.2% | 12.4% | 12.4% | 4.8% | 4.8% |
| 2040 | 7.6% | 0.1%<br>(0.0% - 2.6%) | 11.3% | 0.7%<br>(0.2% - 4.2%) | 7.2% | 0.1%<br>(0.0% - 2.4%) | 4.1% | 0.6%<br>(0.2% - 1.8%) |
| 2060 | 4.3% | 0.1%<br>(0.0% - 0.7%) | 8.1% | 0.5%<br>(0.2% - 2.2%) | 3.8% | 0.0%<br>(0.0% - 0.5%) | 4.4% | 0.5%<br>(0.1% - 1.7%) |
| 2080 | 3.4% | 0.1%<br>(0.0% - 0.6%) | 7.6% | 0.5%<br>(0.1% - 2.0%) | 2.8% | 0.0%<br>(0.0% - 0.4%) | 4.8% | 0.5%<br>(0.1% - 1.6%) |
| 2100 | 3.2% | 0.1%<br>(0.0% - 0.6%) | 7.4% | 0.5%<br>(0.1% - 2.0%) | 2.7% | 0.0%<br>(0.0% - 0.4%) | 4.7% | 0.5%<br>(0.1% - 1.6%) |

Notes: MD = Major Depression. Parenthetical estimates reflect results under optimistic and pessimistic policy effect estimates under a nicotine product standard. See supplement for details. Current smoking is defined as having smoked at least 100 cigarettes in one's lifetime and smoking at all within the past year, which includes those who recently quit and are likely to relapse.

The policy was associated with 190,000 (70,000 discounted) premature deaths averted for those with MD and 1.5 million premature deaths averted (940,000 discounted) for those without MD from 2027-2100 (Table 2). Those with MD experienced a greater relative change in deaths averted from the policy compared to those without depression (2.5% vs. 0.7%). The decrease in premature deaths was similarly reflected in LYG, with 74.7 million (22.9 million discounted) for the total population. Under the product standard, people with current MD lose 3.0 million (780,000 discounted) life years, reflecting the decreased number of people with MD under the product standard in general, as reductions in smoking lead to reductions in depression across the population.

**Table 2.** Simulated Cumulative Deaths Averted and Life Years Gained, 2027-2100, by Major Depression Status in status quo and nicotine product standard (policy) scenarios es

| Outcome | Overall Population |  | Population with MD |  | Population without MD |  |
| --- | --- | --- | --- | --- | --- | --- |
|  | Status Quo | Policy | Status Quo | Policy | Status Quo | Policy |
| Cumulative Deaths (millions) | 222.6 | 221.0<br>(220.9 - 221.3) | 7.7 | 7.5<br>(7.5 - 7.6) | 214.9 | 213.4<br>(213.4 - 213.7) |
| Cumulative Premature Deaths Averted (millions) |  | 1.7<br>(1.3 - 1.7) |  | 0.2<br>(0.1 - 0.2) |  | 1.5<br>(1.2 - 1.5) |
| Discounted Cumulative Premature Deaths Averted (millions) |  | 1.0<br>(0.7 - 1.1) |  | 0.1<br>(0.0 - 0.1) |  | 0.9<br>(0.6 - 1.0) |
| Relative % Change in Premature Deaths Averted |  | 0.7%<br>(0.6% - 0.8%) |  | 2.5%<br>(1.9% - 2.6%) |  | 0.7%<br>(0.5% - 0.7%) |
| Cumulative Life Years (millions) | 22877.7 | 22952.3<br>(22929.8 - 22957.2) | 2031.6 | 2028.7<br>(2027.5 - 2029.6) | 20846.0 | 20923.7<br>(20900.2 - 20929.7) |
| Cumulative Life Years Gained (millions) |  | 74.7<br>(52.1 - 79.5) |  | -3.0<br>(-4.1 - -2.1) |  | 77.6<br>(54.2 - 83.6) |
| Discounted Cumulative Life Years Gained (millions) |  | 22.9<br>(14.5 - 24.8) |  | -0.8<br>(-1.2 - -0.5) |  | 23.6<br>(15.0 - 26.0) |

Notes: MD = Major Depression. Population deaths were calculated by scaling annual deaths in the simulated population to total annual U.S. deaths. Population life years were calculated by scaling each simulated birth cohort to the number of actual births in the US. Discounted estimates reflect a 3% discount rate applied starting in 2025. The negative number of life years gained in the population with MD reflects the decreased number of people with MD under a nicotine product standard, as smoking reductions also decrease MD episodes. Relative % change in Premature Deaths Averted is calculated as deaths averted (policy-status quo)/ status quo. Parenthetical estimates reflect results under optimistic and pessimistic policy effect estimates under a nicotine product standard. Note, numbers in parenthetical estimates are ordered low to high for readability, regardless of optimistic or pessimistic effects.

### Cost Outcomes

The nicotine product standard was estimated to be highly cost-effective from a healthcare perspective: $13,378 in medical costs per QALY gained (Table 3). Societal costs are higher at $53,734/QALY, but still well below the $100,000 ICER threshold. At the US population level, the product standard is projected to increase healthcare costs by $296 billion from 2027–2100, while also increasing overall productivity by $266 billion and consumer spending by $1.16 trillion.

**Table 3.** Incremental Cost Effectiveness Ratios and Simulated U.S. Cumulative Costs, 2027-2100, under a nicotine product standard.

| **Outcome**<br>**ICER** | **Total Population** |
| --- | --- |
| Medical Costs per QALY | 13,378<br>(12,226 - 13,681) |
| Societal Costs per QALY | 53,734<br>(52,065 - 53,724) |
| Medical Costs per LY | 12,964<br>(11,982 - 13,359) |
| Societal Costs per LY | 52,073<br>(51,025 - 52,463) |
| **U.S. Level Outcomes**<br>**(billions)** |  |
| Cumulative US Medical Costs | 296<br>(194 - 297) |
| Cumulative US Consumer Expenditures | 1,159<br>(737 - 1,260) |
| Cumulative US Productivity Losses | -266<br>(-292 - -168) |
| Cumulative US Societal Costs | 1,190<br>(762 - 1,266) |

Notes: ICER = incremental cost-effectiveness ratio. QALY= Quality-Adjusted Life Year. LY= Life Year. U.S. level cost outcomes were calculated by multiplying the corresponding ICER to the discounted cumulative Life Years Gained (LYG). Negative productivity losses are interpreted as productivity gains.

### Sensitivity Analysis

Across all policy effect parameters evaluated, the impact of a nicotine product standard on smoking cessation confers the most benefit: 92.5% of the MPRPM can be achieved through the effects of the policy on cessation alone (eTable 5). When varying the excess mortality risk associated with vaping, more premature deaths were averted and life years gained when higher excess mortality risk due to vaping is assumed: 75.4 million LYG at 15% excess risk vs. 73.2 million at 0% excess risk (eTable 6). If MD incidence probabilities were to return to their lower pre-2016 level (eFigure 2), the model estimated health care cost savings under a nicotine product standard, highlighting the impact of MD on overall healthcare costs. (eTable 7). However, changing assumptions about future trends in MD had negligible impact on smoking prevalence or cumulative premature deaths averted by 2100. See Supplement for details.

## Discussion

This is the first study to quantify the impact of a nicotine product standard on the population by mental health status, explicitly incorporating dynamics between smoking, vaping, and depression. We estimated that the product standard would be associated with 1.7 (1.3-1.7) million premature deaths averted and 74.7 (52.1-79.5) million life years gained from 2027-2100. A nicotine product standard would greatly accelerate declines in smoking, bringing smoking prevalence among people with and without MD to near-zero by 2040. In absolute terms, it would not exacerbate smoking and mortality disparities between people with and without MD.

Our study also demonstrates that concerns about the harms of a nicotine product standard for people with mental health conditions are unfounded. Reducing nicotine in cigarettes could even have modest mental health benefits: our model showed that a nicotine standard would be associated with a 0.06% percentage point decrease in MD prevalence, equivalent to 8.5 million fewer adults with MD by 2100. These results are consistent with evaluations of other tobacco policies that have been shown to affect mental health, e.g., smoke-free bans in workplaces and homes have been associated with reduced risk of depression.^38,39^ Given the growing recognition of the nation’s mental health crisis and demand from the public to address it, policies that have the potential to improve mental well-being strengthen the case for intervention and may also be more favorably received by decision-makers.

This study is also the first to quantify healthcare costs, productivity gains, and consumer expenditures associated with a nicotine product standard. Our model does not find healthcare cost savings under the policy; we find the opposite: $296 billion in higher healthcare costs this century. This result is expected and explained by longer life expectancies under the standard—a net positive for society. Though people who never smoked use fewer healthcare resources than people with histories of smoking, older adults use far more healthcare services regardless of smoking status. That said, a nicotine product standard is highly cost-effective. Consider that cancer treatments with much higher ICERs are still recommended and covered by insurance policies.^40^ Under a willingness to pay threshold of $100,000/QALY, our finding of $13,378/QALY gained makes a nicotine product standard an incredible bargain, like other policies geared towards chronic disease prevention. Longer, healthier lives under a nicotine product standard also benefits the economy; our study estimates $266 billion dollar increase in productivity this century.

Furthermore, Our model results for the total population are largely consistent with projections reported by the FDA.^1,25^ Our estimates of projected smoking prevalence and life-years gained by 2100 are similar (0.1% vs 0.2% and 74.7 vs. 76.4 million), while estimates of premature deaths averted are more conservative but have overlapping uncertainty intervals (1.7 (1.3, 1.7) vs. 4.3 (1.6, 4.6) million).

There are limitations in our analysis. First, we lack information on the real-world effects of a nicotine product standard on smoking and vaping. The policy effects in our model were based on expert elicitation, rather than cohort or RCT data, and thus may not reflect the actual effects of a nicotine product standard. Second, we assumed that the policy effects of a nicotine product standard were the same for those with and without MD. Third, we did not model the potential effects of an illicit market for NNCs.^40,41^ Consumer access to illicit NNCs would dampen the overall magnitude of public health benefits under a nicotine standard (fewer people would quit smoking), but would not change the direction of our results. Fourth, we only considered cigarettes and e-cigarettes, not other tobacco products such as cigars. For a nicotine product standard to be successful, nicotine levels would also need to be reduced for cigars so that consumers do not have a harmful and highly addictive combustible alternative on the market. We also use MEPS data, which has been noted to underreport health care use and thus may underestimate costs.^41^ Finally, we did not include other costs such as those incurred to enforce compliance. However, our model may have underestimated improvements to health as the potential benefits incurred by reductions in second and third-hand smoke exposure and environmental harms.

Although a federal nicotine product standard remains under consideration, significant workforce reductions at the FDA Center for Tobacco Products Office of Regulations are likely to delay further steps.^42^ However, states have the legal authority to pursue nicotine reduction policies through state-level sales restrictions.^43^ Doing so would increase the availability of real-world evidence on the effects of reduced nicotine cigarettes on tobacco use behavior. Any nicotine reduction policy is certain to face tobacco industry litigation. Its success will depend in part on the extent to which industry arguments influence public sentiment: ninety percent of adults believe there is a mental health crisis in the US today,^44^ so claims that a nicotine product standard could harm people with mental health conditions must be debunked. ^7,8^

Our results suggest that a nicotine reduction strategy is a cost-effective intervention that will benefit people with and without MD through substantial reductions in tobacco product use and mortality. Timely implementation at the state or federal level could help Americans live longer and healthier lives, address tobacco use disparities by mental health status, and benefit the economy through increases in overall productivity.

## Funding

Research reported in this publication was supported by the National Institute on Drug Abuse (K01DA056424). Dr. Tam and Dr. Skolnick are also supported by the National Cancer Institute (U01CA253858, U54CA229974). Dr. Brouwer is supported by the National Cancer Institute (U54CA229974).

## Disclaimer

The opinions expressed in this article are the authors’ own and do not reflect the views of the National Institutes of Health, the Department of Health and Human Services or the US government.

## Additional Contributions

We thank Drs. Stephen Higgins, Rafael Meza, Suchitra Krishnan-Sarin for their guidance during this study. We also wish to thank Drs. Susan Busch and A. David Paltiel for their feedback on this manuscript.

## Data Availability

Data from the National Surveys on Drug Use and Health (NSDUH) are publicly available online.

https://www.samhsa.gov/data/data-we-collect/nsduh-national-survey-drug-use-and-health/datafiles

## Notes

### Competing Interest Statement

The authors have declared no competing interest.

### Author Declarations

Data from the National Surveys on Drug Use and Health (NSDUH) are publicly available online. https://www.samhsa.gov/data/data-we-collect/nsduh-national-survey-drug-use-and-health/datafiles

### Summary of Updates

Author name was changed from Charlene Cheng to formal name Chia-ling Cheng

