## Supplement for "Health, Equity, and Economic Impacts of a Nicotine Product Standard in the United States for People With and Without Major Depression"

**Supplemental Online Content**

### eMethods

#### MDSV Model

Figure 1 displays the model structure and transitions. We allow for annual transitions between health states with only one state change per year, except for product switching which is defined as switching from exclusive smoking to exclusive e-cigarette use. The model was calibrated to NSDUH data using the following definitions:

Never smoking (N) is defined as having smoked less than 100 cigarettes in a lifetime, current smoking (C) is defined as having smoked 100 or more cigarettes and smoked at all in the past year, and former smoking (F) is defined as having smoked 100 or more cigarettes and not smoked in the past year.

Never e-cigarette use is defined as having never vaped (O), current e-cigarette use is defined as having vaped in the past 30 days (E), and former e-cigarette use is defined as having vaped but not in the past 30 days (Q).

Never MD is defined as individuals reporting no lifetime history of major depressive episode (H), current MD is defined as a major depressive episode in the past year (D), and former or recovered MD is defined as a lifetime major depressive episode but not in the past year (R). Depression is defined differently for different sources. For example, some studies use a formal diagnosis of major depressive disorder and some use self-reported depression. The NSDUH uses self-reported Major Depressive Episode which is defined as at least five of the following nine symptoms for a two-week or longer period: depressed mood; diminished interest in activities; changes in weight or appetite; insomnia or hypersomnia; psychomotor symptoms; fatigue; feeling worthless; problems thinking; thoughts of death or suicide. This is based on DSM-5 criteria.^9^

The MDSV model was developed in R using the Decision Analysis in R for Technologies in Health (DARTH) work group coding and microsimulation modeling framework.^1,2^ All R code used for this study will be available at <https://github.com/jamietam/mdse-microsim> upon publication.

#### Model Calibration

The development of a simulation model typically involves the use of parameters with unknown values that are not readily available from the literature or other data sources. For such parameters of unknown value, we estimate those values by calibrating model outputs to the observed survey data. This process adjusts the parameter values to ensure that the model outputs align with our calibration targets, specifically the NSDUH 2005-2023 prevalence data. We estimated the relative risk of MD among those who smoke, the relative risk of quitting smoking among those with MD, and the relative risk of initiating vaping among those with MD, among others (See eTable 1). Transition probabilities of smoking, depression, and e-cigarette use were obtained from a variety of data sources and were subsequently scaled to fit NSDUH data through calibration. Calibration targets included tobacco use prevalences (smoking, e-cigarette, dual use), prevalence of MD, and MD prevalences by tobacco use status by gender. Calibration for the MDSV model was conducted by searching for sets of parameter values that maximize the log likelihood value, a goodness-of-fit measure that evaluates the likelihood of the observed data being produced by the model. The model then performs simulations using these best-fit parameter values.

#### Costs

Healthcare costs assigned to each individual by age, smoking status, and depression status were based on data from the Medical Expenditure Panel Survey (MEPS). Health care costs by smoking status were obtained from an analysis of linked National Health Interview Survey (NHIS) and Medical Expenditure Panel Survey (MEPS) using the Manning two-part model to estimate medical expenditure by smoking status.^3^ Health care costs by depression status were obtained from an analysis by Egede et al. of depression and diabetes using MEPS data from 2004-2011.^4^ We added costs by smoking and by depression to get combined costs. We think this may be conservative because, according to the analysis by Egede et al., the additive effect of diabetes and depression was lower than the estimated joint effect.

#### Utilities

To estimate utilities by depression status, smoking status, and e-cigarette use, we utilized 2022 data from the Behavioral Risk Factor Surveillance System (BRFSS).^5^ Depression status was measured using a self-reported item based on the question: “Has a doctor, nurse, or other health professional ever told you that you had a depressive disorder (including depression, major depression, dysthymia, or minor depression)?” Responses were categorized dichotomously (yes/no). Smoking status was also self-reported and classified into three categories: current smoker, former smoker, and never smoker. The classification was based on responses to two questions: “Have you smoked at least 100 cigarettes in your entire life?” and “Do you now smoke cigarettes every day, some days, or not at all?” E-cigarette use was self-reported and categorized into two groups: current users and non-current users. This was assessed through the question: “Would you say you have never used e-cigarettes or other electronic vaping products in your entire life or now use them every day, use them some days, or used them in the past but do not currently use them at all?”

Health utility was derived from self-reported healthy days based on the question: “During the past 30 days, for about how many days did poor physical or mental health keep you from doing your usual activities, such as self-care, work, or recreation?” We stratified the number of healthy days by depression status, smoking status, and e-cigarette use. We then converted the reported number of healthy days into a health utility score ranging from 0 to 1 using the estimation method developed by Jia and Lubetkin.^6^

#### Nicotine Product Standard Policy Effects

Experts were asked what they thought the effects would be of a nicotine product standard on initiation, cessation, dual use, product switching, and vaping initiation among those who would have smoked without the standard. As in the FDA analysis, we use the median values from the expert elicitation in our main scenario, with the 5^th^ and 95^th^ percentile values from the expert elicitation as upper and lower bound estimates. These effects of the product standard applied in our model are available in eTable 4. Furthermore, eFigure 1 shows the pathway/transition each effect acts on. Numbers in eFigure 1 correspond to numbers in eTable 4.

#### Sensitivity Analysis Results

eTable 5 shows cumulative mortality reductions when policy effects are applied one at a time. The impact of a nicotine product standard on smoking cessation confers the most benefit: 92.5% of the MPRPM is achieved through the effects of the policy on cessation. Reductions to smoking initiation account for 22.0% of the MPRPM, while product switching from cigarettes to e-cigarettes achieves 24.9%

eTable 6 shows mortality reductions under the main policy scenario (10% excess mortality risk due to vaping) and under 0% and 15% excess vaping mortality risk. More premature deaths are averted and life years gained when higher excess mortality risk due to vaping is assumed; 73.2 vs. 75.4 million LYG for 0% and 15% excess risk, respectively.

eFigure 2 displays MD prevalence under status quo scenario with incidence constant from 2016-2100 vs returning to pre-2016 levels starting in 2023. Assuming a lower depression incidence (pre-2016 levels) results in health care cost savings of 478.2 billion due to many fewer depressed individuals (eTable 7). However, smoking prevalence and cumulative premature deaths averted by 2100 are similar between the two depression incidence scenarios. The cumulative premature deaths averted is 1.65, assuming the incidence is constant from 2016 to 2100 and 1.66 assuming incidence returns to pre-2016 levels starting in 2023.

### eFigures

#### eFigure 1. Overview of policy effects in MDSV model.

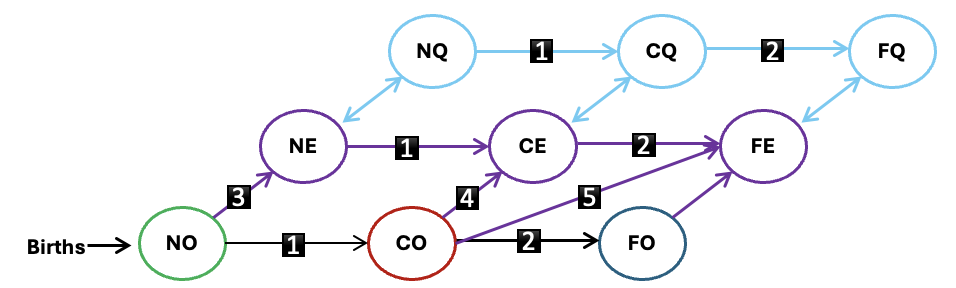

N = Never smoked, C = Current smoking. F = Former smoking, O = Never e-cigarette use, E = Current e-cigarette use, Q = Former e-cigarette use. Numbers refer to policy effects on tobacco use transition probabilities as specified in eTable 4.

eFigure 2. MD prevalences under status quo and nicotine product standard scenarios.

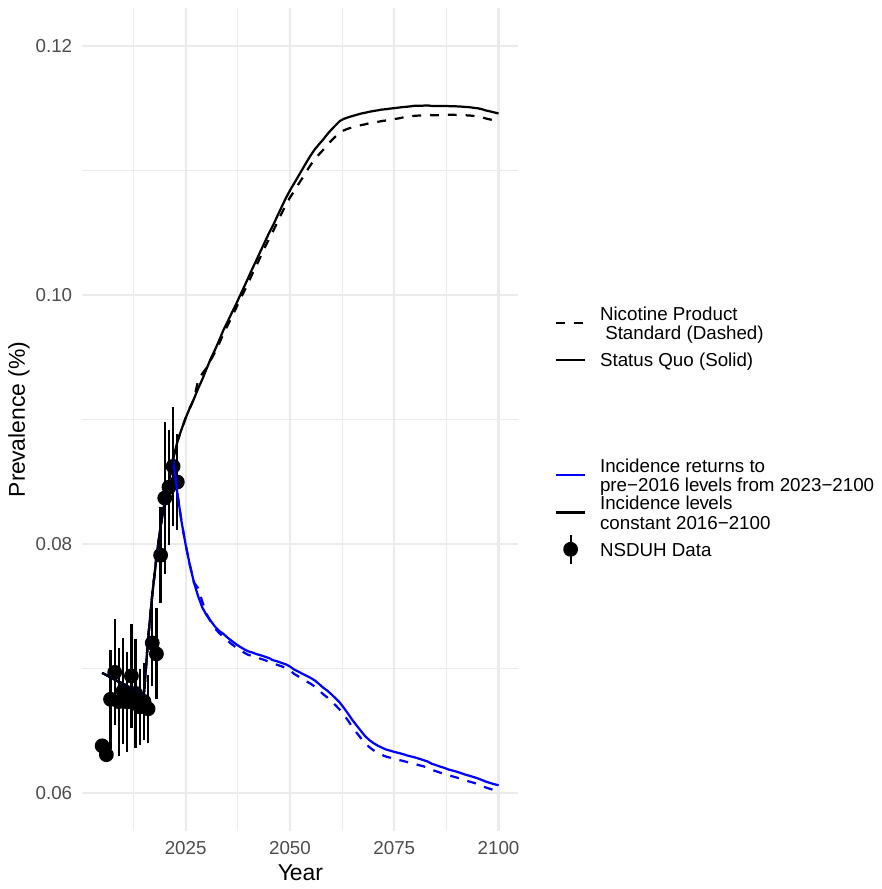

Notes: MD = major depression. The solid lines represent the status quo scenarios, and the dashed lines are the Nicotine Product Standard scenarios. The blue lines represent scenarios with projected depression incidence set at 2016 levels, and the black lines represent scenarios with projected depression incidence set at 2022 levels. Dots are NSDUH data points used to calibrate the model. Vertical lines on dots are confidence intervals.

#### eFigure 3. Major depressive episode prevalence among adults, NSDUH 2005-2023.

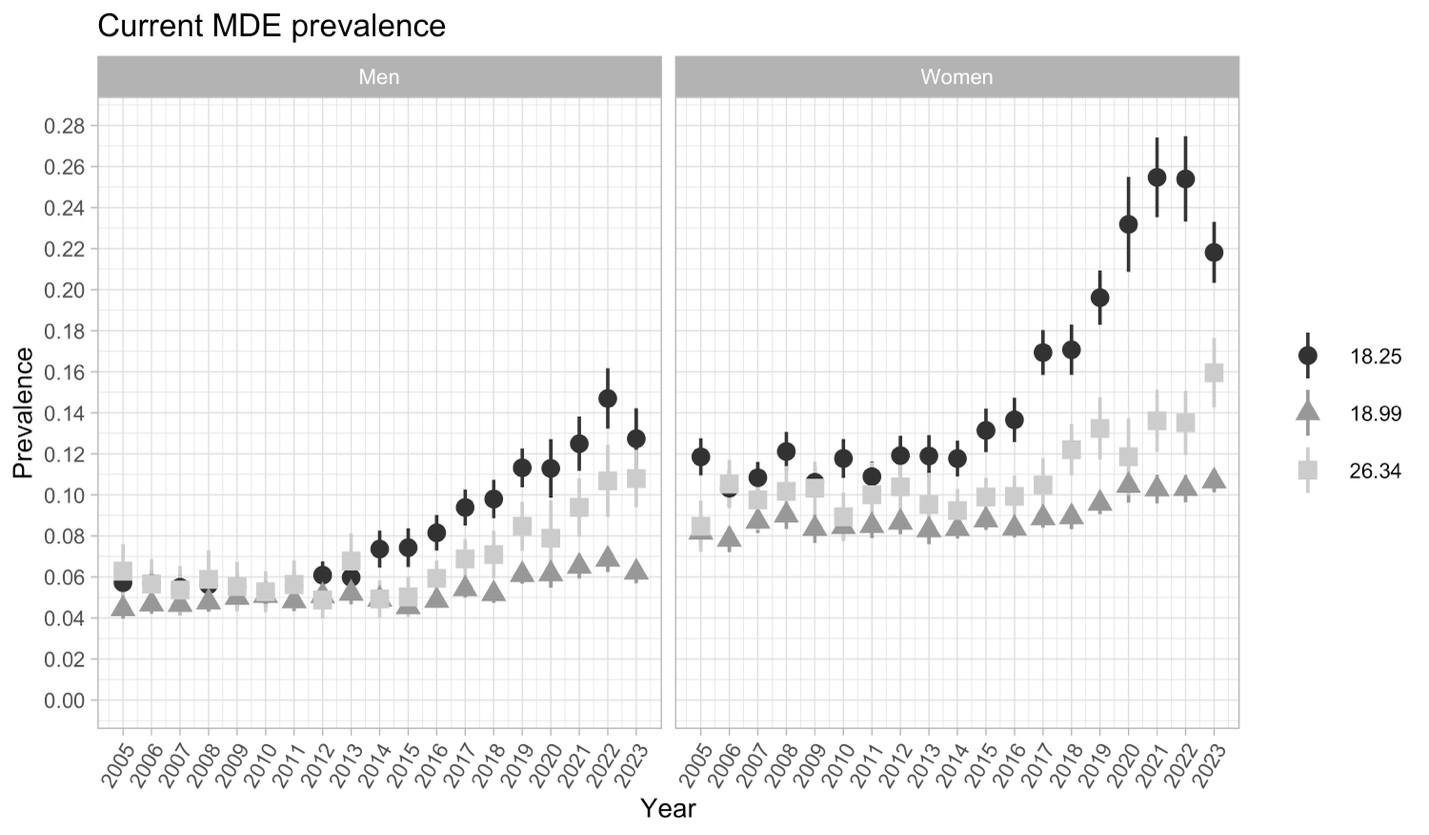

Notes: NSDUH = National Survey on Drug Use and Health. A major depressive episode is defined as a two-week or longer period during which the respondent reports at least 5 of 9 depressive symptoms in the past year.

### eTables

#### eTable 1. MDSV parameters and data sources.

| Parameter | Data Source |
| --- | --- |
| Probability of MD incidence (p.HD) | The incidence of first onset of MD for women ages 22+ and men ages 29+ is based on data from the Baltimore-ECA study.^7^ Incidence of first onset MD for women aged 12 to 21 and men ages 12 to 28 was unavailable from the Baltimore-ECA study, and where therefore estimated using cubic natural splines.^8^ To capture rising levels of MD among young adults (eFigure 3), scaling factors (s.HD) were applied to increase MD incidence probabilities by age group starting in 2016 (2-4X increase). Given uncertainty regarding future MD incidence among young people, sensitivity analysis explored whether allowing MD incidence to return to their pre-2016 levels would change model outcomes (See eTable 7). |
| Probability of recovering from MD (p.DR) | Set to be constant at 0.173 for ages 12-64. Previously calculated in Tam 2020 from the Baltimore-ECA cohort study.^8,9^ Because of low MD prevalence among older adults, the probability for ages 65+ for men and women is estimated through calibration. |
| Probability of MD recurrence (p.RD) | Set to be constant at 0.058. Previously calculated in Tam 2020 from the Baltimore-ECA cohort study.^8,9^ |
| Probability of smoking initiation (p.NC) | Gender, age, and birth cohort specific initiation probabilities were obtained from the Cancer Intervention and Surveillance Modeling Network (CISNET) Lung Working Group analysis of National Health Interview Surveys (NHIS) ^10^ These are then calibrated to the NSDUH 2005-2023 data using scaling factors by age group (s.NC). Initiation probabilities are held constant for the most recent birth cohort (2008) going forward. |
| Probability of smoking cessation. (p.CF) | Gender, age, and birth cohort specific cessation probabilities were obtained from the CISNET Lung Working Group analysis of National Health Interview Surveys (NHIS).^10^ These cessation probabilities are age, sex, and birth-cohort specific and reflect permanent cessation, with no relapse back to smoking. These are calibrated to the NSDUH 2005-2023 data using scaling factors by age group (s.CF), with cessation probabilities held constant for the 2010 birth cohort going forward. |
| Transition probabilities between smoking and e-cigarette use states | See eTable 3 below and Figures S3 in the Supplements to Brouwer et al.^11–13^ |
| Relative risk of MD incidence among people who currently smoke (rr.CH.CD) | Gender specific risks estimated through calibration. |
| Relative risk of MD recurrence among people who currently smoke (rr.CR.CD) | Gender specific risks estimated through calibration. |
| Relative risk of smoking cessation among people with MD (rr.CD.FD) | Gender specific risks estimated through calibration. |
| Relative risk of vaping initiation among people with MD (rr.OD.ED) | Gender specific risks estimated through calibration. |
| Probability of death by smoking status (p.NX, p. CX, p.FX.ysq). | Gender, age, and birth cohort specific death probabilities were obtained from the CISNET Lung Working Group using the relative risks of all-cause mortality described in the Supplement of Tam 2021.^14^ Mortality among those who formerly smoked varies by years since quitting.^15,16^ |

#### eTable 2. Vaping transitions and data sources.

| Years | 2013-2014^13^ | 2015-2016^12^ | 2017-2019 ^12^ | 2020-2021^17^ |
| --- | --- | --- | --- | --- |
| Ages | Never use to exclusive vaping | | | |
| 12-14 | 0.006 | | 0.013 | |
| 15-17 | 0.026 | | 0.054 | |
| 18-24 | 0.01 | 0.01 | 0.018 | 0.017 |
| 25-34 | 0.003 | 0.002 | 0.002 | 0.002 |
| 35-54 | 0.001 | 0.001 | 0 | 0 |
| 55-90 | 0.001 | 0 | 0 | 0 |
|  | Exclusive smoking to dual use | | | |
| 12-14 | 0.16 | | 0.419 | |
| 15-17 | 0.152 | | 0.223 | |
| 18-24 | 0.073 | 0.069 | 0.102 | 0.151 |
| 25-34 | 0.055 | 0.051 | 0.052 | 0.068 |
| 35-54 | 0.039 | 0.034 | 0.034 | 0.023 |
| 55-90 | 0.02 | 0.015 | 0.013 | 0.023 |
|  | Non-current to exclusive vaping | | | |
| 12-14 | 0.324 | | 0.162 | |
| 15-17 | 0.182 | | 0.333 | |
| 18-24 | 0.046 | 0.057 | 0.118 | 0.089 |
| 25-34 | 0.018 | 0.017 | 0.019 | 0.029 |
| 35-54 | 0.009 | 0.008 | 0.007 | 0.003 |
| 55-90 | 0.002 | 0.002 | 0.002 | 0.003 |
|  | Exclusive vaping to non-use | | | |
| 12-14 | 0.46 | | 0.393 | |
| 15-17 | 0.402 | | 0.235 | |
| 18-24 | 0.313 | 0.329 | 0.193 | 0.204 |
| 25-34 | 0.199 | 0.194 | 0.137 | 0.244 |
| 35-54 | 0.187 | 0.206 | 0.113 | 0.129 |
| 55-90 | 0.111 | 0.101 | 0.114 | 0.129 |
|  | Dual use to exclusive smoking | | | |
| 12-14 | 0.352 | | 0.008 | |
| 15-17 | 0.377 | | 0.184 | |
| 18-24 | 0.413 | 0.435 | 0.152 | 0.18 |
| 25-34 | 0.457 | 0.433 | 0.23 | 0.225 |
| 35-54 | 0.451 | 0.462 | 0.265 | 0.323 |
| 55-90 | 0.44 | 0.435 | 0.371 | 0.323 |
|  | Exclusive Smoking to exclusive vaping | | | |
| 12-14 | 0.089 | | 0.105 | |
| 15-17 | 0.04 | | 0.058 | |
| 18-24 | 0.018 | 0.019 | 0.032 | 0.045 |
| 25-34 | 0.015 | 0.013 | 0.017 | 0.012 |
| 35-54 | 0.008, | 0.006, | 0.007 | 0.003 |
| 55-90 | 0.007 | 0.006 | 0.004 | 0.003 |
| Notes: All numbers shown are rates which are converted to probabilities for the model. See Figures S3 in the Supplements to Brouwer et al. ^11–13^ | | | | |

#### eTable 3. Economic evaluation data inputs.

| Parameter | Source |
| --- | --- |
| Consumer Expenditures (Consumption of non-healthcare goods). | U.S. Bureau of Labor Statistics. Consumer Expenditure Survey: Calendar Year Tables. ^18^ |
| Medical/Healthcare costs | Health care costs by smoking status were obtained from an analysis of linked National Health Interview Survey (NHIS) and Medical Expenditure Panel Survey (MEPS) using the Manning two-part model to estimated Medical expenditure by smoking status.^3^  Health care costs by depression status were obtained from an analysis by Egede et al. of depression and diabetes using MEPS data from 2004-2011.^4^ We added costs by smoking and by depression to get combined costs. We think this may be conservative because in the Egede et al analysis, the additive effect of diabetes and depression was lower than the estimated joint effect. This analysis produced results for 2015 dollars and thus we inflated to 2023 dollars to apply in our model using the Price Index for health care services. |
| Productivity (Total mean wage by age group, both sexes combined) | U.S. Census Bureau, Current Population Survey, 2023 Annual Social and Economic Supplement (CPS ASEC).^19^ The fringe rate in 2023 was 31% among civilian workers.^20^ Fringe benefits were included in calculation of wages. |
| Utilities (QALY) accounting for smoking, e-cigarette use, and depression | 2022 Behavioral Risk Factor Surveillance System (BRFSS) data was used to calculate number of healthy days by depression status, smoking, and e-cigarette use. We then converted the reported number of healthy days into a health utility score ranging from 0 to 1 using the estimation method developed by Jia and Lubetkin.^6^ |

#### eTable 4. Nicotine product standard policy effects.

| Effect on tobacco use behavior | 1st year of implementation | Subsequent year |
| --- | --- | --- |
| 1. Smoking initiation: % reduction in initiation | 63% (38-83%) | 65% (39-85%) |
| 2. Smoking cessation: proportion that will quit | 36% (11-61%) | 34% (11-56%) |
| 3. Dual use: proportion that will dual use cigarettes and e-cigarettes | 61% (25-90%) | 51% (19-82%) |
| 4. Product switching: proportion of individuals that will switch completely from cigarettes to e-cigarettes | 56% (22-84%) | 58% (25-85%) |
| 5. E-cigarette initiation: proportion of individuals deterred from smoking that will initiate vaping | 50% (21-72%) | 50% (20-75%) |

Notes: [Provide citation here]

#### eTable 5. Sensitivity analysis 1: most impactful policy effect.

| Parameter | Cumulative Premature Deaths Averted by 2100 (millions) | % of MPRPM |
| --- | --- | --- |
| MPRPM | 1.73 | - |
| Main Scenario | 1.65 | 95.4% |
| 1. Smoking initiation: % reduction in initiation | 0.38 | 22.0% |
| 2. Smoking cessation: proportion that will quit | 1.60 | 92.5% |
| 3. Dual use: proportion that will dual use cigarettes and e-cigarettes | 0.04 | 2.3% |
| 4. Product switching: proportion of individuals that will switch completely from cigarettes to e-cigarettes | 0.43 | 24.9% |
| 5. E-cigarette initiation: proportion of individuals deterred from smoking that will initiate vaping | 0.38 | 22.0% |
| Notes: Smoking Attributable Deaths Averted are in millions. We simulated a Maximum Potential Reduction in Premature Mortality (MPRPM) scenario, which assumes all individuals quit smoking and no one initiates smoking. Thus the percentage of the MPRPM explain the benefit of each effect to an ideal scenario in which individuals stop smoking. | | |

#### eTable 6. Sensitivity Analysis 2: E-cigarette Mortality Effects

| Excess mortality risk due to Vaping | Cumulative Premature Deaths Averted (millions) | Cumulative Life Years Gained  (millions) |
| --- | --- | --- |
| 10% (Main Scenario) | 1.65 | 74.65 |
| 0% (Lower estimate) | 1.60 | 73.21 |
| 15% (Upper estimate) | 1.69 | 75.35 |
| Notes: Excess mortality risk is equivalent to 10% of the excess mortality of current smoking compared to never smoking. | | |

#### eTable 7. Sensitivity Analysis 3: Major Depression Incidence Scenarios

| MD incidence | Current smoking prevalence among people with MD under main effect by 2100 | Cumulative Premature Deaths Averted  (millions) | Cumulative Health care costs  (billions) |
| --- | --- | --- | --- |
| Assumes annual MD incidence probabilities remain constant from 2016-2100 (main scenario) | 0.1% | 1.65 | 296.3 |
| Assumes annual MD incidence probabilities return to pre-2016 levels from 2023-2100 | 0.0% | 1.66 | -478.2 |
| Notes: Premature deaths averted are in millions and health care costs are in billions. | | | |

20.	Employer Costs for Employee Compensation – March 2023. Accessed March 23, 2025. www.bls.gov/ebs.
